# Risk of lung cancer among welders and flame cutters: A Systematic Review and Meta-Analysis

**DOI:** 10.1101/2022.08.29.22279357

**Authors:** Saptorshi Gupta

## Abstract

**Background and Aim:** The process of flame-cutting and welding is believed to highly hazardous for workers involved in related industries. The study aims to provide a comprehensive quantitative effect of the risk of lung cancer due to exposure to welding fumes.

**Methods:** A systematic review was conducted to extract published estimates of odd’s ratio of the association of lung cancer and exposure to welding fumes, till 2022. Studies were extracted from the PubMed database with clearly defined inclusion and exclusion criteria. Studies from all parts of the world were considered. A meta-analysis with random effects model was conducted resulting in formation of forest plot, influence analysis, sensitivity analysis and checking for publication bias using a funnel plot.

**Results:** The meta-analysis yielded an OR of 1.256 (95% CI 1.055-1.496), with a moderately high heterogeneity between the studies (*I*^2^ = 68.78% ; *τ*^2^ = 0.039; *Q* = 36.115(*p* < 0.001)). The sensitivity and influence analysis confirmed the absence of highly influential studies that may have led to potentially distorted outcomes. The funnel plot showed no evidence of publication bias among the studies included in this analysis.

**Conclusion:** As the association of lung cancer and occupational hazards from exposure to welding fumes is certain, there is a need to control and regulate industrial activities that involve welding and flame cutting. Already, restrictions on safe levels of fume in workplace is in operation.

## Introduction

The process of flame cutting, which is primarily a Thermo-Chemical Process. was first introduced in 1901 by Thomas Fletcher. The mechanism requires pure oxygen and a source of intense heat, often extending beyond 1000 degrees Fahrenheit. This process is capable of cutting ranging from sheet metal thickness to 100-inch material. The appropriate part of the material cut is raised to ignition temperature by an oxy-fuel gas flame. Also referred to as oxy-fuel cutting, the process is primarily implemented for separating and shaping steel components.

In the case of oxy-fuel welding, a welding torch is used to weld metals by heating two pieces of the metal, heated to a particularly high temperature, thus producing a molten pool. The concern revolving around carcinogenicity of welding fumes have been long researched about (IARC, 1990; Simonato et al, 1991). Welding fumes, however, have been identified as highly carcinogenic for humans (IARC, 2017). In the process of welding, a compound concoction of fumes may develop from base metal, fluxes, shielding gases, electrode and surface coatings. Different welding processes generate different combinations of particulate matter and metals (Kendzia et al, 2013; Weiss et al, 2013). Hence, welders may be exposed to work-place related carcinogens including ionizing radiation, asbestos and silica.

Studies have found association of exposure to welding fumes with immune suppression (Grigg et al., 2017; Marongiu et al., 2016), oxidative stress marker (Han et al., 2005; Hoffmeyer et al 2012) and systemic inflammation (Kim et al., 2005; Shen et al., 2018; Wang et al., 2005). Work related occupational diseases include occupational asthma or pneumoconiosis. In addition, welding in confined spaces is fatally dangerous. Worldwide, about 11 million welders and 110 million additional workers are exposed to fumes emitted from welding (Guha et al., 2017).

Industries like shipbuilding involve welding of large amounts of mild steel. Such an occupational setting is linked to high levels of exposure to respirable particles (Lehnert et al, 2012; Weiss et al, 2013). A lot of epidemiological studies exist linking exposure to welding and lung cancer. Some of the early studies, although not consistently statistically significant, especially the cohort studies including welders, have shown an increase in risk of lung cancer (Beaumont and Weiss, 1981; Fletcher and Ades, 1984). Eventually, more studies have been conducted focusing on specific forms of welding fumes like gas and arc welding fumes (Becker, 1999; Jockel 1998), stainless steel or mild steel welding (Simonato et al, 1990; Steenland et al, 1991; Moulin et al, 1993) and shipyard welding (Steenland et al, 1991; Moulin et al, 1993). These studies, however, have shown varied results without an explicit pattern of association. Yet, some studies have reported definite higher risk of lung cancer in welders with longer and higher cumulative exposure (Matrat et al. 2016; Siew et al. 2008; Sorensen et al. 2007; t Mannetje et al. 2012). The extensive use of asbestos in welding operations could play the role of a confounder in association with welding fumes (Puntoni et al, 1979; Palmer and Eaton, 2001).

Thus, to establish a definite association between exposure to welding fumes and lung cancer, a meta-analysis on the past studies have been performed.

## Methods

The present systematic review and meta-analysis aimed to calculate the odds of higher risk of lung cancer due to exposure to occupational hazards arising from welding and flame cutting based on the Preferred Reporting Items for Systematic Reviews and Meta-Analysis (PRISMA) guidelines 2020 (Page *et al*., 2021).

### Literature search

The electronic literature search was conducted using the PubMed database for locating and accessing studies related to lung cancer and its association to different forms of occupational hazards prevalent at workplaces. Studies were limited to those which used the case-control design and were published till the year 2022. The keywords used for this search were ‘lung cancer’, ‘occupational/industrial exposure’, ‘welding fumes’, ‘fumes’, ‘arc fumes’ and ‘case-control’. Only those studies that could be retrieved as full length, freely available articles and were published in English were considered for this study. The reference list of such articles was also screened for additional support.

### Inclusion and exclusion criteria

Studies pertaining to association of various forms of occupational hazards, as exposure variables, to the incidence of lung cancer (outcome variable) were included for analysis. Only those studies which allowed extraction of data such that a 2*2 contingency table could be created to estimate odds ratio at 95% confidence interval (CI) were taken into account for this meta-analysis. In short, the given selection criteria were applied for the selection of studies:

- Studies must have a case-control design
- Studies must have reported a 2*2 contingency table for recalculation the odds ratios and their respective 95% Confidence Intervals (CIs) of cancers related to occupational exposures, separately.

Studies that reported mixed effects of carcinogens along with prevalence of smoking were excluded if the effects of carcinogens could not be shown separately. Additionally, full-length reports that were unable to be accessed in English were also excluded.

A flow chart showing a simple explanation of the study process along with the number of studies included and excluded have been shown in the results section (Figure 1).

**Figure 1:**
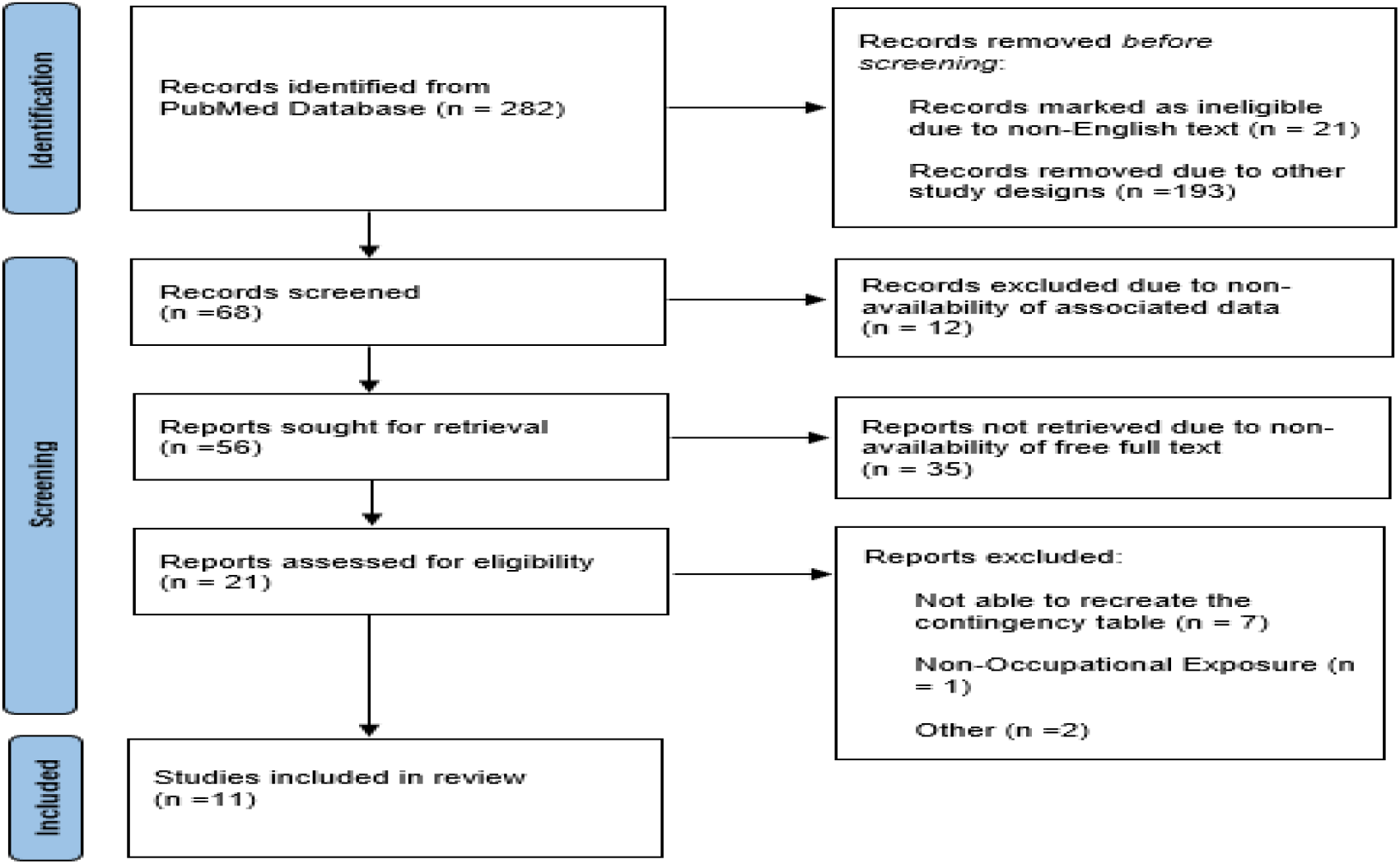
PRISMA flow diagram showing the selection process of studies included to understand the carcinogenic impacts of welding fumes exposure on lungs.

### Data extraction

The Consolidated Standards of Reporting Trials (CONSORT) and Strengthening the Reporting of Observational studies in Epidemiology (STROBE) was reviewed for data extraction. From each of the studies the following information was abstracted: Last name of the first author, Year of Publication of the study, Location, duration and industry type involving the study population, Number of cases exposed to occupational carcinogens, Total number of controls, Number of controls exposed to occupational carcinogens, Total number of controls, Carcinogens that the subjects were exposed to.

### Statistical analysis

The R software (meta and metafor packages) was used for analysis, creation of statistical plots and describing individual studies and pooled ORs.

For each of study, 2*2 contingency tables were created to recalculate odds ratio and the 95% confidence interval (CI) by following the standard procedure. The overall odds ratio and its 95% confidence interval was calculated based on the heterogeneity of the analysis. For heterogeneity of >50%, the random effects model was used. Statistical significance was set at a p value of less than 5%.

A p-curve was plotted to check for substantial evidential values present. Eventually, contribution of heterogeneity and effect size was checked through Baujat plot and influence analysis. A sensitivity analysis was performed to check the effect of removal of outliers on the effect size and heterogeneity.

### Assessment of heterogeneity

Heterogeneity of the studies that were taken for analysis in this review were analyzed using three types of heterogeneity measures – Cochran’s Q statistic, Higgin’s & Thompson’s *I*^2^ and the Tau-squared statistic, with a standard alpha value of 0.05 used for statistical significance.

The Cochran’s Q-statistic was used to understand the difference between the observed effect sizes (OR) and the fixed-effect model estimate of the effect size. Although this measure is not used to assess the extent of heterogeneity, a low p-value (p<0.05) associated with the Q-statistic has been used to confirm the presence of heterogeneity.

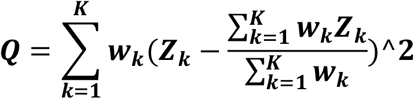

Where k is the individual study, K is indicative of all studies in the meta-analysis and *Z*_*k*_ is the estimated effect of k with a variance of 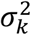, and *w*_*k*_ is the individual weight of the study.

The *I*^2^ statistic is used to understand the percentage of variability in the effect sizes which is not caused by sampling error (Higgins and Thompson, 2002). *I*^2^ values less than 50% were considered as low level of heterogeneity and those above 50% were considered high.

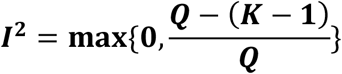

Since *I*^2^ statistic is heavily dependent on the precision of the studies (Rucker et al., 2008; Borenstein et al., 2017), it tends to 100% as the number of studies included becomes large, thus reducing the sampling error. Hence, as another method of heterogeneity check the *τ*^2^ statistic was used to understand the degree of between-study variance in the meta-analysis. It is an estimate of the extent of dispersion of the true effect sizes.

### Bias

Funnel plot at 90%, 95% and 99% confidence intervals were plotted to check for a possible publication bias.

## Results

### Studies included

The PubMed search generated 282 studies based on the keywords ‘lung cancer’, ‘flame cutters’ and ‘welding fumes exposure’. A total of 21 studies were marked as ineligible due to non-English text. Upon inclusion of the ‘case-control’ keyword, 193 studies were excluded due to other study designs. Among the remaining 68 studies, 12 were eliminated due to non-availability of associated data. The resultant 56 studies were sought for retrieval and 35 among them were not retrieved due to non-availability of free full text. 21 publications were assessed for eligibility among which 7 were excluded as it was not able to recreate contingency tables, 1 were excluded due to non-occupational exposures and 2 were excluded due to other reasons. After going through exclusion and inclusion criteria and making the final set of selections, 11 studies were selected for analysis. The strategy for identification of studies is presented in figure 3. These studies have all looked at the odds of the incidence of lung cancer due to occupational exposure to welding and flame cutting.

The key characteristics of the studies are given in Table 1. All the studies in this meta-analysis are case controlled studies. The studies included in this analysis were highly heterogenous in terms of region. 3 studies were selected from the United States of America (Breslow *et al*., 1954; Zahm *et al*., 1989; Morabia *et al*., 1992), 2 from Canada (Vallières *et al*., 2012), 1 from South America (Pezzotto and Poletto, 1999), 1 multicentric study involving Central and Eastern Europe and USA (‘T Mannetje *et al*., 2012) and 4 from Europe which included 2 studies from Italy (Buiatti *et al*., 1985; Ronco et al, 1988), 1 study from France (Benhamou, Benhamou and Flamant, 1988) and 1 study from Germany (Pesch *et al*., 2019).

**Table 1:** Table showing the study characteristics of papers included to understand the association between incidence of lung cancer and exposure to welding fumes.

| Sl No | Study Name | Study Period | Region | Baseline Population | Objective of Study | Result |
| --- | --- | --- | --- | --- | --- | --- |
| 1 | Breslow et al | 1949-1952 | California, USA | 518 cases and 518 controls | To determine whether any additional occupation apart from a preselected few deserved intensive investigation. | Very high odds were linked to lung cancer as a result of welding. |
| 2 | Buiatti et al | 1981-1983 | Florence, Italy | 376 cases and 892 controls | Investigation of occupational risk factors for both men and women. | High risk of lung cancer was found among welders and bricklayers using firebrick and other refractory materials. |
| 3 | Benhamou et al | 1976-1980 | France | 1625 cases and 3091 controls | To make precise estimate of the proportion of cancers attributable to occupation. | An excess risk of lung cancer was observed for welders among a few selective professions, after making adjustment for cigarette exposure. |
| 4 | Ronco et al | 1976-1980 | Italy | 126 cases and 384 controls | Understanding the risk of welding as a contributing factor of lung cancer. | Welders showed an elevated odds ratio in relation to lung cancer. |
| 5 | Zahm et al | 1980-1985 | USA | 4431 cases and 11326 controls | To evaluate the relationship between lung cancer histologic types and occupation, adjusted for smoking. | Elevated risk of lung cancer was reported among welders. |
| 6 | Morabia et al | 1980-1989 | USA | 1793 cases and 3228 controls | To estimate the risk of lung cancer attributable to occupational factors not due to tobacco. | Occupational factors play an important role in the incidence of lung cancer independently of cigarette smoking. |
| 7 | Pezzotto & Poletto | 1980-1986 | Rosario, Argentina | 367 cases and 576 controls | To evaluate exposure in working environment after controlling factors of cigarette smoking. | Occupational exposures are found to partly account for high lung cancer mortality rate among male residents. |
| 8 | Vallieres et al Study 1 | 1979-1986 | Montreal | 857 cases and 1066 controls | To investigate relationships between occupation exposure to gas and arc welding fumes and the risk of lung cancer. | There was no significant excess risk of lung cancer among heavy smokers due to welding fumes. However, among light smokers, an excess risk was detected, linked to welding fumes. |
| 9 | Vallieres et al Study 2 | 1996-2000 | Montreal | 736 cases and 894 controls |  |  |
| 10 | Mannetje et al | 1998-2001 | 6 Central and Eastern European countries and | 3403 cases and 3670 controls | To elucidate the extent to which confounding by smoking and asbestos in driving the association and | A duration response association was noted for mild steel welding without chromium exposure. |
|  |  |  | Liverpool, United Kingdom |  | to evaluate the role of welding related exposure. |  |
| 11 | Pesch et al | 1988-1996 | Germany | 3418 cases and 3488 controls | To investigate the risk of lung cancer after exposure to welding fumes, chromium and nickel. | Welding fumes, Cr (VI) and nickel are believed to contribute independently to the excess lung cancer risk associated with welding. |

### Meta Analysis Results

#### Heterogeneity

The studies included in this meta-analysis has reported significantly high heterogeneity (Table 2).

**Table 2:** Heterogeneity of studies for each individual carcinogen.

| Statistic | Value | Confidence Interval |
| --- | --- | --- |
| Higgin's & Thompson's $I^2$ | 72.3% | [49.1%, 84.9%] |
| Tau-squared statistic | 0.0524 | [0.0153,0.9600] |

( $Q=36.12$ , $p<0.0001$ )

**Table 3:**

| Test | Value | P value |
| --- | --- | --- |
| Egger et al (1997) | 0.97 | 0.3590 |
| Peters et al. (2006) | 0.06 | 0.9511 |

#### Effect Size

The meta-analysis has been reported in three forms – 4 studies reporting association of only gas welding fumes and lung cancer (Figure 3), 4 reporting the association of only arc welding fumes (Figure 4) and lung cancer and finally a pooled result from all the 11 studies on the incidence of lung cancer (Figure 2) due to occupational exposure to welding and flame cutting yielding OR of 1.28 (95% CI 1.05-1.55), with a moderately high heterogeneity between the studies (*I*^2^ = 72.3% ; *τ*^2^ = 0.05; *Q* = 36.12(*p* < 0.001)). The meta-analysis summary showed 1.2 times higher odds for incidence of lung cancer if exposed to flames emitted from welding in occupational settings.

**Figure 2:**
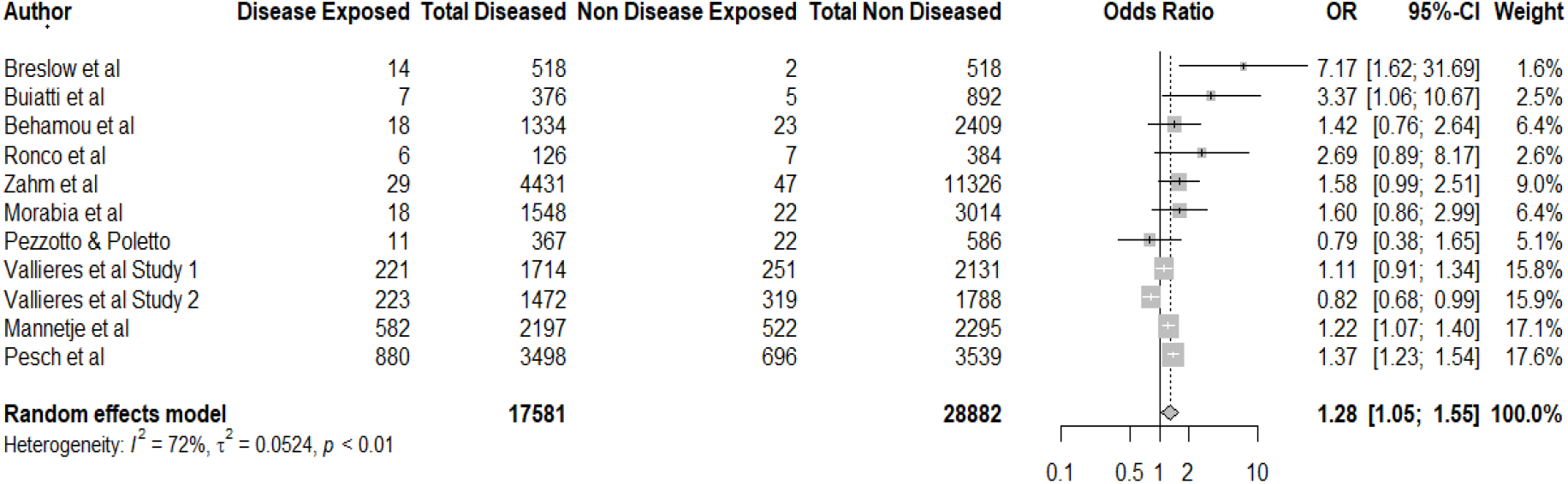
Forest Plot showing the pooled association between exposure to welding fumes and lung cancer.

**Figure 3:**
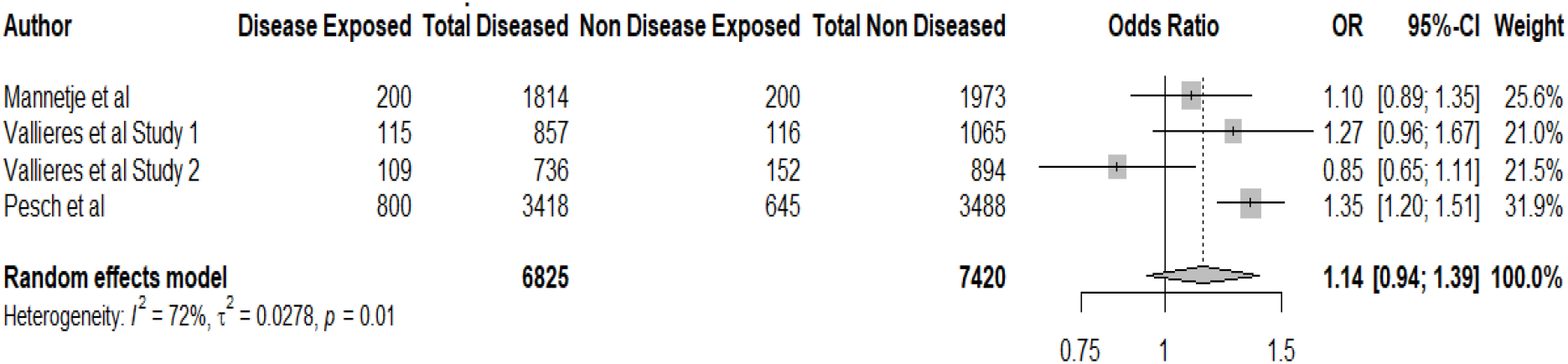
Forest Plot showing the pooled association between only welding fumes exposure and lung cancer.

**Figure 4:**
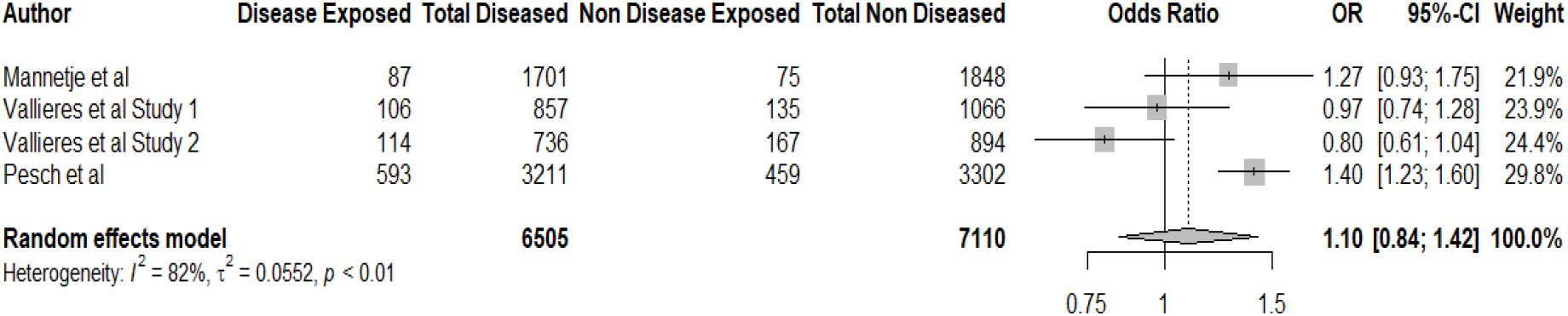
Forest Plot showing the pooled association between arc fume exposure and lung cancer.

Figure 3 and 4 show forest plot of the individual association of gas welding fumes and arc fumes on lung cancer. The association of exposure to only welding fumes or only arc fumes to the development of lung cancer did not yield significant outcomes overall. Among the studies included, only one study showed positive association with statistical significance (Pesch et al, 2019).

In the pooled meta-analysis including all the studies selected, four studies reported its association of lung cancer incidence with occupational exposure to all possible fumes emitted during welding and flame cutting (Breslow et al, 1953; Buiatti et al, 1985; Mannetjee et al, 2012; Pesch et al, 2019). The ORs of these studies ranged from 1.11 to 7.17. The remaining studies did not yield statistically significant result of its association (Behamou et al, 1988; Ronco et al, 1988; Zahm et al, 1989; Morabia et al 1992; Pezzotto and Poletto, 1999; Vallieres et al, 2012). This analysis concludes that the exposure to welding fumes and flame cutting is linked to about 1.28 times to the risk of incidence of lung cancer.

#### P-Curve

The p-curve (Figure 5) presents substantial evidential value present in the analysis. Among the 11 studies provided, 5 (45.45%) of them included into the analysis have p<0.05 and 3(2.27%) of them have a p value of less than 0.025. There is, thus, conclusive evidence that the set consists mostly of studies with true effects as there are more p-values between 0 and 0.01 than between 0.04 and 0.05.

**Figure 5:**
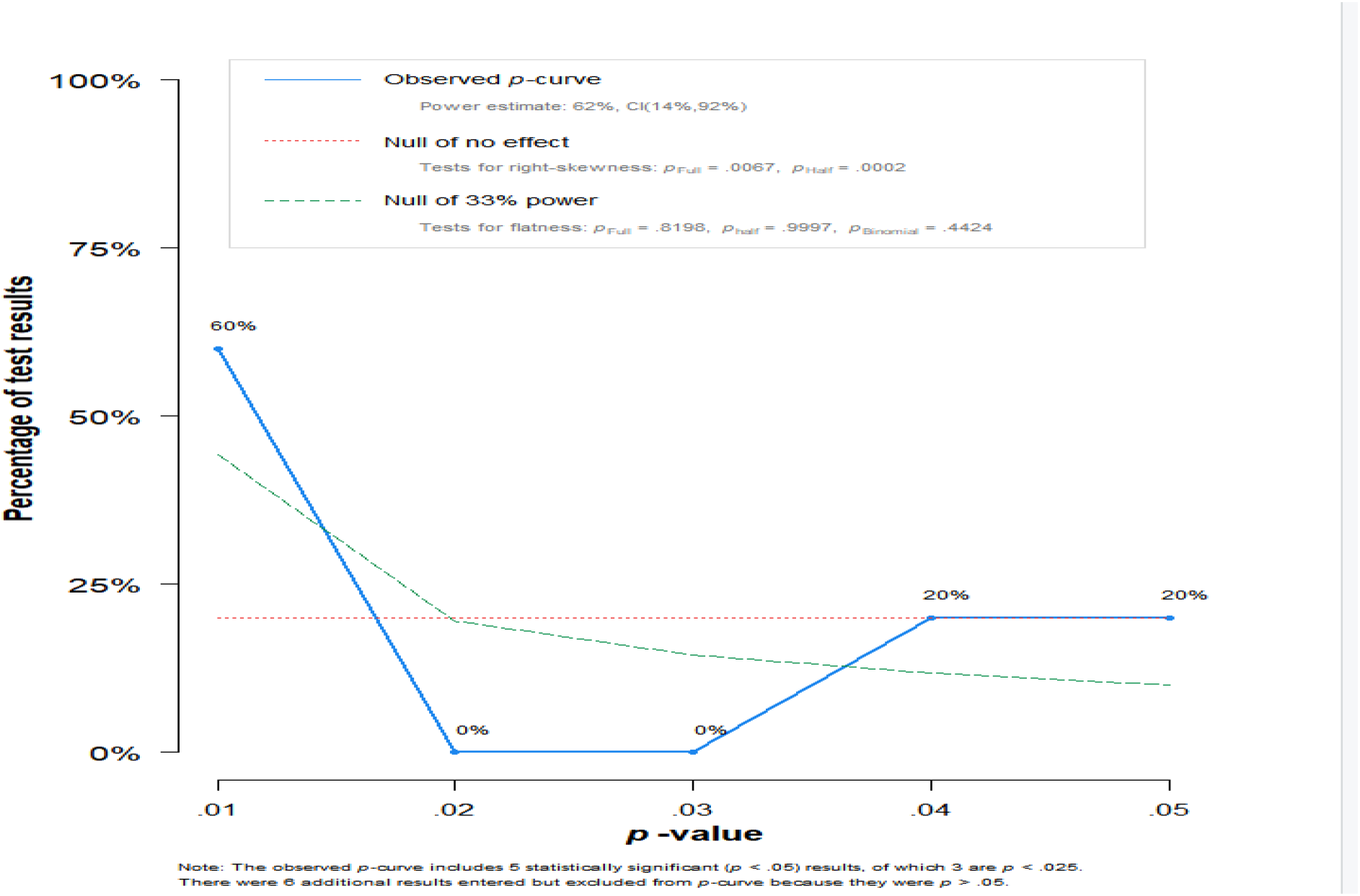
p-Curve showing evidential value of the studies.

#### Contribution to Heterogeneity and Influence Analysis

The diagnostic Baujat plot (Figure 6) detected Vallieres et al Study 2, that had excessively contributed to the heterogeneity of the meta-analysis. Pesch et al and Vallieres et al Study 2 had significantly higher contribution to the overall effect size.

**Figure 6:**
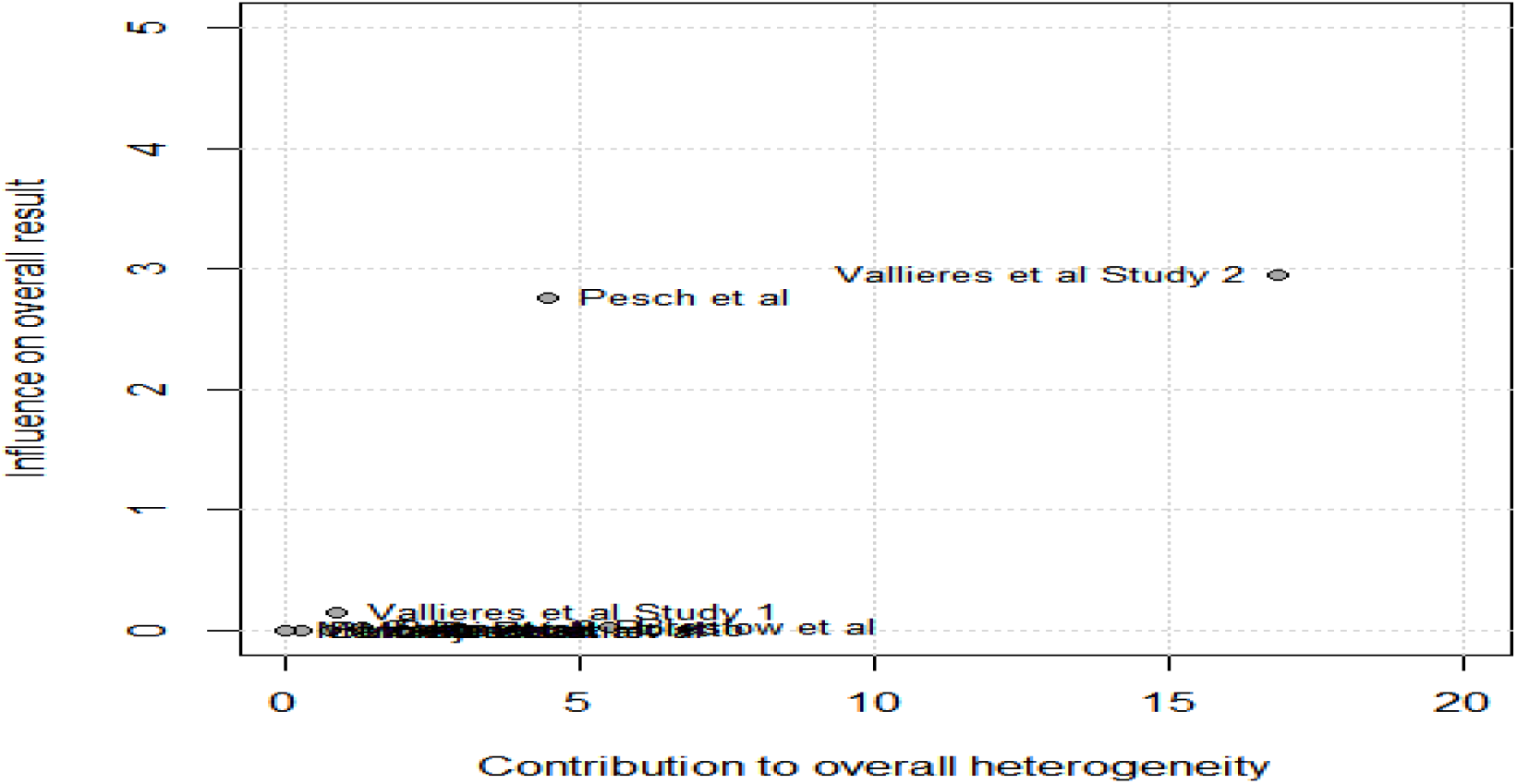
Baujat plat showing outliers among the studies included.

The influence diagnostic plots (Figure 7) state how well the studies fit into the meta-analysis model. As is represented in the graph, none of the studies are displayed in red (indicative of high influence), in any of the diagnostics. Thus, none of the studies had a significant distortionary effect on the pooled effect estimate. ’

**Figure 7:**
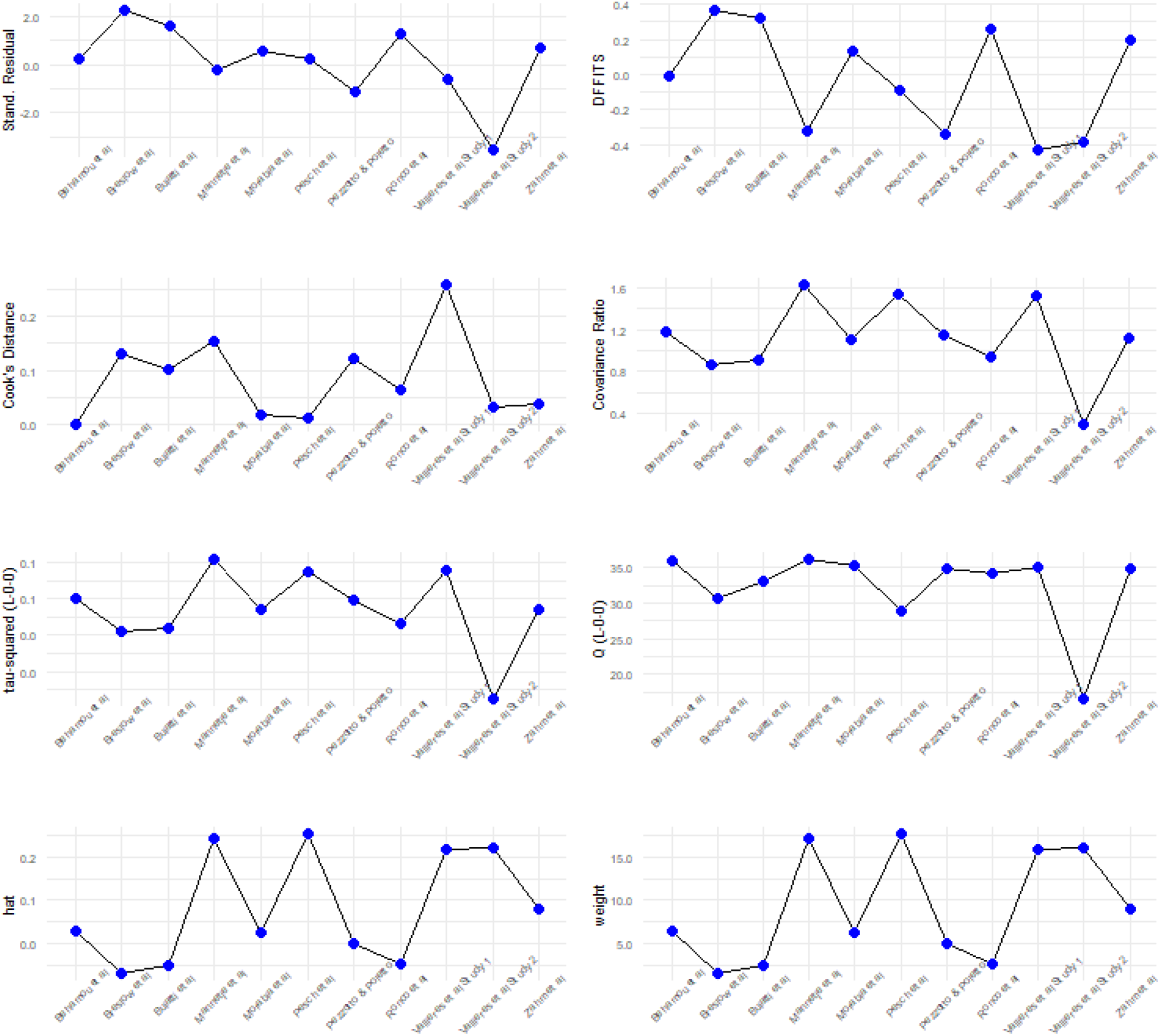
Influence Analysis.

#### Sensitivity Analysis

The Baujat plot identified a potential outlier in the study Vallieres et al Study 2. Upon removal of the outliers (Figure 8), the pooled odd’s ratio is 1.28 (95% C.I. 1.16-1.42) which stays significant, indicating that even after removal of the outliers, the overall conclusion from the analysis remains unchanged. The heterogeneity of the studies has decreased considerably (*I*^2^ = 30.0% [95% *C. I*. 0.0% − 67.6%]).

**Figure 8:**
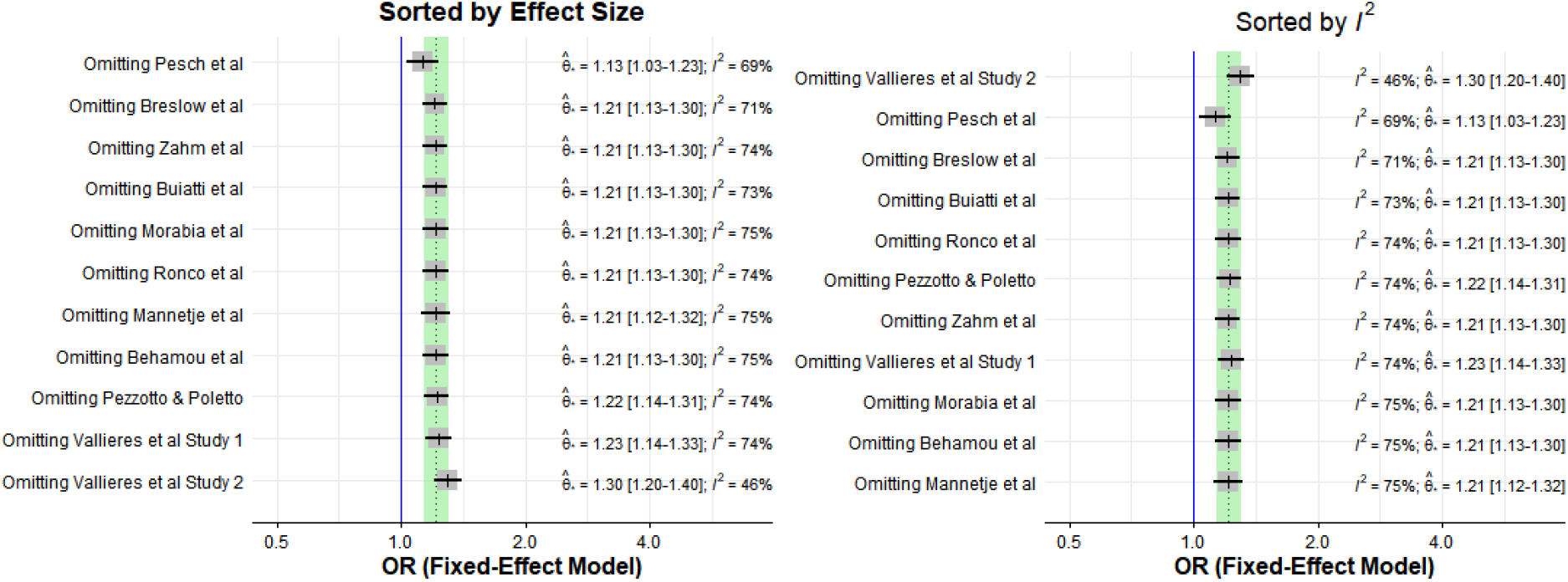
Sensitivity Analysis.

**Figure 9:**
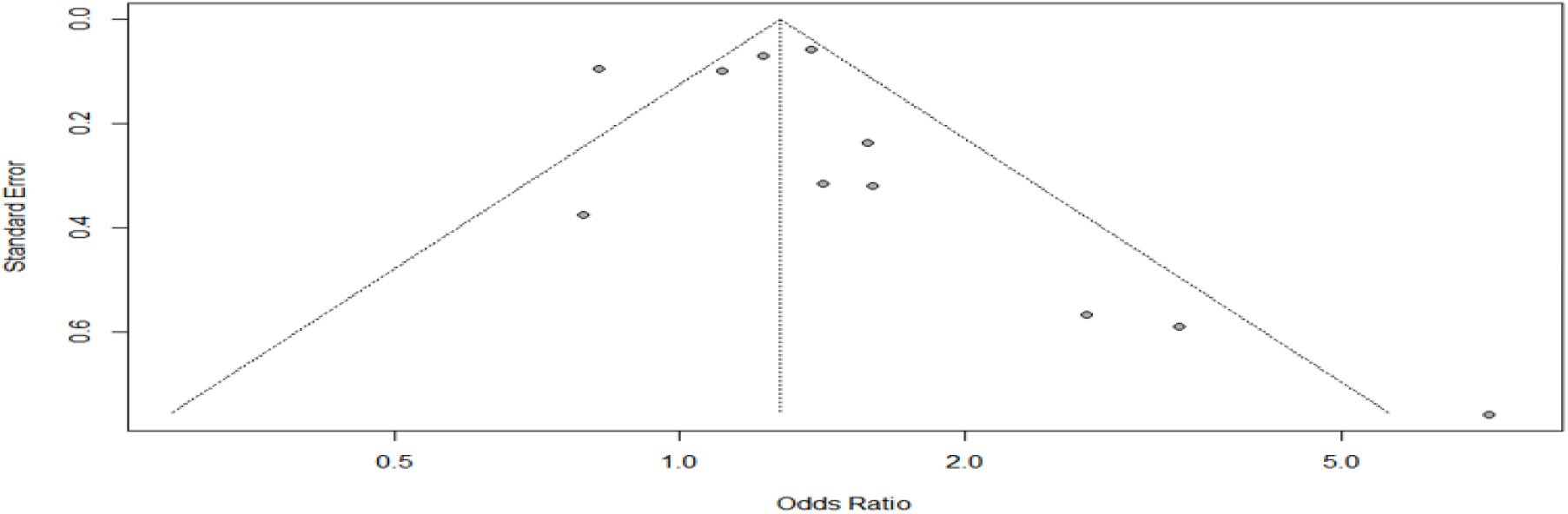
Funnel Plot for studies associating lung cancer and exposure to welding fumes.

**Figure 10:**
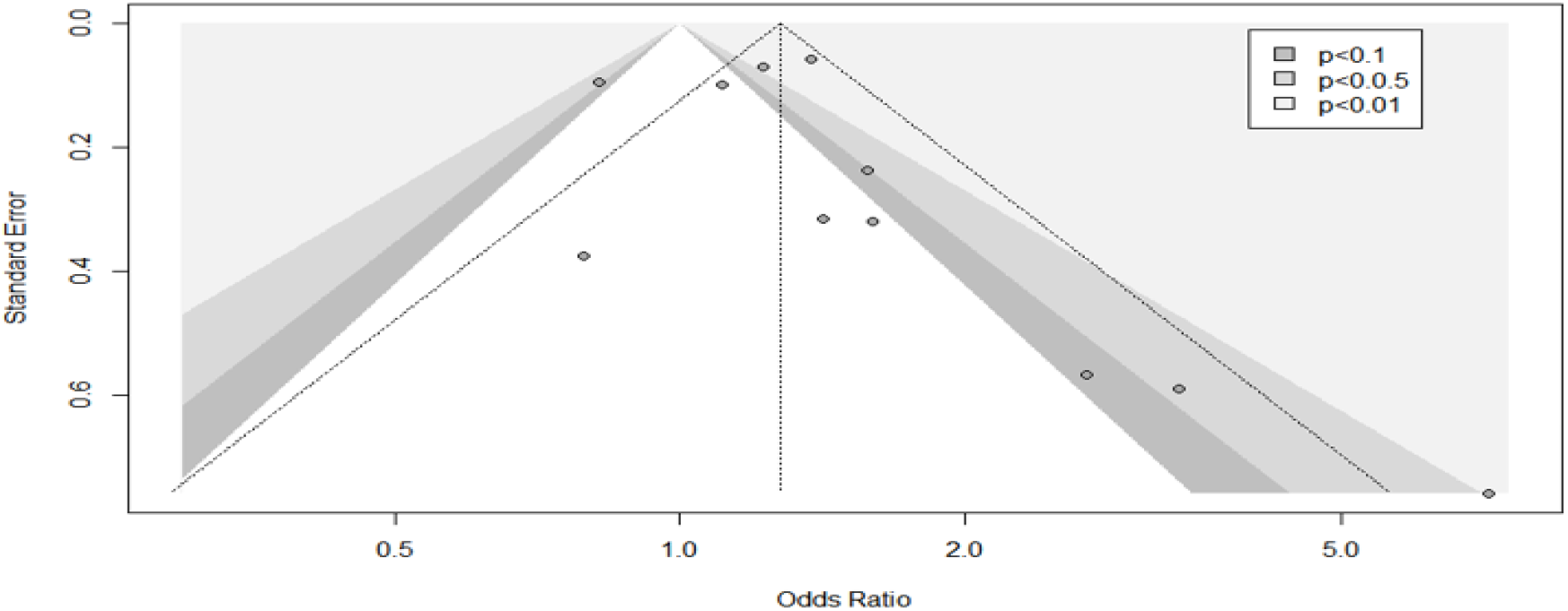
Contour-Enhance Funnel Plot.

#### Bias

Funnel plots are devised to understand the presence of publication bias in studies. A contour-enhanced funnel plot is used to understand the statistical significance that are overlaid on the funnel plot (Peters et al, 2008

The use of Egger’s test (1997) and Peter’s Test (2006) confirms the absence of asymmetry in the meta-analysis.

The presence of equal share of studies in the right-hand side of the vertical line suggests that fairly similar number of studies have been published reporting effects due to occupational exposures and with no association.

## Discussion

This systematic review of association between occupational exposure to welding fumes and lung cancer involved analysis of 11 papers that had evaluated this association in different populations. The intensity of risks was calculated on the basis of information extracted on exposure and non-exposure of cases and controls. Based on the level of heterogeneity (*I*^2^ > 50%), the analysis was based on binary random effects model. The pooled results of the meta-analysis were satisfactorily significant (p=0.010) to provide evidence for increased risk of cancer incidence among welders and flame cutters. As has been mentioned in section 4.3, the review included study from a number of different regions. This can be attributed to the high heterogeneity detected in the report. Additionally, working conditions, socio-economic situation and other attributes may have been a cause for this.

Reviews involving case-control and cohort studies have been performed in the past to evaluate the effect of welding fumes in the incidence of lung cancer (Sjogren et al, 1994; Kendzia et al, 2013; Falcone, 2018; Honaryar et al, 2019). The study by Sjogren was a meta-analysis of five studies of stainless-steel welders and their association of the occurrence of lung cancer. The analysis yielding a pooled relative risk estimate of 1.94 suggested a causal relation between exposure to stainless steel welding and lung cancer, having taken into account the confounding effects of smoking habits and asbestos exposure (Sjogren et al, 1994). Similarly, the meta-analysis by Honaryar performed a meta-analysis on 37 non-overlapping study populations and compared the incidence of lung cancer among never-exposed and ever-exposed to welding fumes. The study found increase in risk of lung cancer due to exposure to welding fumes, regardless of the type of steel welded, welding methods and exposure to confounders like asbestos or tobacco smoking (Honaryar et al, 2019). Results from Kendzia et al., that included data from male lung cancers and controls from 16 studies in Europe, Canada, China and New Zealand, revealed an increase in the odds of incidence of lung cancer among welders. As intuitive as it may seem, the odds ratio was found to increase for longer durations of welding (Kendzia et al, 2013). The study by Falcone also supported the fact that welders using mild or stainless steel were at an increased risk for lung cancer. This meta-analysis is consistent with the previous studies conducted. Although, the association of the individual effects of welding fumes or arc gas fumes was not significant, the outcome can be attributed to the low number of studies that could be accommodated. In this regard, this calls for further research to investigate the effects of welding fumes or arc fumes only, without any confounders.

## Conclusion

As the association of lung cancer and occupational hazards from exposure to welding fumes is certain, there is a need to control and regulate industrial activities that involve welding and flame cutting. Already, restrictions on safe levels of fume in workplace is in operation. Much of the regulatory framework is intended towards protection of health of workers through maintenance of exposure to fumes and gases with a predetermined limit. In the United Kingdom, the limits are referred to as Workplace Exposure Limits (WELs). Two time periods – short and long, are used to check the exposure time limits and prevent health hazards. Similarly, in the United States, the Occupational Safety and Health Administration (OSHA), offers guidelines to help welders reduce their exposure levels to welding fumes and other hazardous gases. OSHA’s Hazard Communication standard is required by employers to provide information and training to workers and elucidate the guidelines for working with hazardous materials. In Ireland, the Control of Substances Hazardous to Health Regulations (COSHH) prevents exposure to substances hazardous to health, lest the workers fall ill. The control of welding fumes is deemed satisfactory if principles of control are applied and exposure to substances within fumes are below the Workplace Exposure Limit (WEL). There is a pressing need for such stringent regulatory policies and safety standards in all industries across the world.

## Supporting information

COI Disclosure

## Data Availability

All data produced in the present work are contained in the manuscript.

## References

Beaumont JJ, Weiss NS. Lung cancer among welders. J. Occup. Med. 1981; 23:839–844.

Becker N. Cancer mortality among arc welders exposed to fumes containing chromium and nickel – results of a third follow-up: 1989–1995. J. Occup. Environ. Med. 1999; 41:294–303.

Blot WJ, Fraumeni JF (1996) Cancers of the lung and pleura. In: Cancer Epidemiol Prev, Schottenfeld DJF (ed). pp 637–665. Oxford University Press: New-York

Damber L, Larsson LG (1985) Underground mining, smoking, and lung cancer: a case-control study in the iron ore municipalities in northern Sweden. J Natl Cancer Inst 74: 1207 – 1213

De Matteis S, Consonni D, Bertazzi PA. Exposure to occupational carcinogens and lung cancer risk. Evolution of epidemiological estimates of attributable fraction. Acta Biomed 2008; 79:34–42

Doll R, Peto R. The causes of cancer: quantitative estimates of avoidable risks of cancer in the United States today. J Natl Cancer Inst 1981;66:1191–308

Fletcher AC, Ades A. Lung cancer mortality in a cohort of English foundry workers. Scand. J. Work Environ. Health. 1984; 10:7–16.

Grigg J, Miyashita L, Suri R (2017) Pneumococcal infection of respiratory cells exposed to welding fumes; Role of oxidative stress and HIF-1 alpha. PLoS One 12:e0173569

Guha N et al (2017) Carcinogenicity of welding, molybdenum trioxide, and indium tin oxide. Lancet Oncol 18:581–582

Han SG, Kim Y, Kashon ML, Pack DL, Castranova V, Vallyathan V (2005) Correlates of oxidative stress and free-radical activity in serum from asymptomatic shipyard welders. Am J Respir Crit Care Med 172:1541–1548

Hoffmeyer F et al (2012a) Impact of different welding techniques on biological effect markers in exhaled breath condensate of 58 mild steel welders. J Toxicol Environ Health Part A 75:525–532

IARC Working Group on the Evaluation of Carcinogenic Risks to Humans. Silica, Some Silicates, Coal Dust and Para-Aramid Fibrils. Lyon, France: IARC, 1997.

IARC, A Review of Human Carcinogens; Part C: Arsenic, Metals, Fibres, and Dusts, 2012, pp. 121–145.

IARC, Beryllium, Cadmium, Mercury and Exposures in the Glass Manufacturing Industry, 1993.

IARC, Supplement: Cadmium and Cadmium compounds, 1997.

IARC. IARC monographs on the evaluation of carcinogenic risks to humans, Vol. 49. Chromium, nickel and welding. Lyon: IARC (International Agency for Research on Cancer); 1990. p. 677.

Jockel KH, Ahrens W, Pohlabeln H, Bolmaudorff U, Muller KM. Lung cancer risk and welding – results from a case-control study in Germany. Am. J. Ind. Med. 1998; 33:313–320.

Kendzia, B., Behrens, T., Jöckel, K. H., et al. (2013). Welding and lung cancer in a pooled analysis of case-control studies. American journal of epidemiology, 178(10), 1513–1525. https://doi.org/10.1093/aje/kwt201

Kim JY, Chen JC, Boyce PD, Christiani DC (2005) Exposure to welding fumes is associated with acute systemic inflammatory responses. Occup Environ Med 62:157–163

Lehnert M, Pesch B, Lotz A, et al. Exposure to inhalable, respirable, and ultrafine particles in welding fume. Ann Occup Hyg. 2012;56(5):557–567.

Mannetje A et al (2012) Welding and lung cancer in Central and Eastern Europe and the United Kingdom. Am J Epidemiol 175:706–714

Marongiu A et al (2016) Are welders more at risk of respiratory infections? Findings from a cross-sectional survey and analysis of medical records in shipyard workers: the WELSHIP project. Thorax 71:601–606

Matrat M et al (2016) Welding, a risk factor of lung cancer: the ICARE study. Occup Environ Med 73:254–261

Moher D, Liberati A, Tetzlaff J, Altman DG. (2009). Preferred reporting of items for systematic reviews and meta-analyses; the PRISMA statement BMJ 2009; 339:b2535 doi:10.1136/bmj.b23535

Moulin JJ, Wild P, Haguenoer JM, Faucon D, Degaudemaris R, Mur JM, et al. A mortality study among mild steel and stainless-steel welders. Br. J. Ind. Med. 1993; 50:234–243.

Page, M.J. et al. (2021) ‘The PRISMA 2020 statement: An updated guideline for reporting systematic reviews’, International Journal of Surgery, 88(March). doi:10.1016/j.ijsu.2021.105906.

Palmer WG, Eaton JC. (2001). Effects of welding on health. XI. Miami: American Welding Society.

Pohlabeln, H., Jöckel, K. H., Brüske-Hohlfeld, I., Möhner, M., Ahrens, W., Bolm-Audorff, U., Arhelger, R., Römer, W., Kreienbrock, L., Kreuzer, M., Jahn, I., & Wichmann, H. E. (2000). Lung cancer and exposure to man-made vitreous fibers: results from a pooled case-control study in Germany. American journal of industrial medicine, 37(5), 469–477. https://doi.org/10.1002/(sici)1097-0274(200005)37:5<469::aid-ajim3>3.0.co;2-d

Puntoni R, Vercelli M, Merlo F, Valerio F, Santi L. (1979) Mortality among shipyard workers in Genoa, Italy. Ann. N. Y. Acad. Sci; 330:353–377.

Ronco, G., Ciccone, G., Troia, B., & Vineis, P. (1988). Occupation and lung cancer in two industrialized areas of northern Italy. International Journal of Cancer, 41(3), 354–358. doi:10.1002/ijc.2910410306

Shen S et al (2018) Welding fume exposure is associated with inflammation: a global metabolomics profiling study. Environ Health 17:68

Siew SS, Kauppinen T, Kyyronen P, Heikkila P, Pukkala E (2008) Exposure to iron and welding fumes and the risk of lung cancer. Scand J Work Environ Health 34:444–450

Simonato L, Fletcher AC, Andersen A, Anderson K, Becker N, Chang-Claude J, et al. A historical prospective study of European stainless steel, mild steel, and shipyard welders. Br. J. Ind. Med. 1991; 48:145–154.

Simonato, L., Fletcher, A. C., Andersen, A., Anderson, K., Becker, N., Chang-Claude, J., Ferro, G., Gérin, M., Gray, C. N., & Hansen, K. S. (1991). A historical prospective study of European stainless steel, mild steel, and shipyard welders. British journal of industrial medicine, 48(3), 145–154. https://doi.org/10.1136/oem.48.3.145

Sorensen AR, Thulstrup AM, Hansen J, Ramlau-Hansen CH, Meersohn A, Skytthe A, Bonde JP (2007) Risk of lung cancer according to mild steel and stainless-steel welding. Scand J Work Environ Health 33:379–386

Steenland K, Beaumont J, Elliot L. Lung cancer in mild steel welders. Am. J. Epidemiol. 1991; 133:220–229.

Steenland K, Burnett C, Lalich N, Ward E, Hurrell J (2003) Dying for work: the magnitude of US mortality from selected causes of death associated with occupation. Am J Ind Med 43: 461 – 482

Steenland K, Loomis D, Shy C, et al. Review of occupational lung carcinogens. Am J Ind Med 1996; 29: 474–90.

Wang Z, Neuburg D, Li C, Su L, Kim JY, Chen JC, Christiani DC (2005) Global gene expression profiling in whole-blood samples from individuals exposed to metal fumes. Environ Health Perspect 113:233–241

Weiss T, Pesch B, Lotz A, et al. Levels and predictors of airborne and internal exposure to chromium and nickel among welders—results of the WELDOX study. Int J Hyg Environ Health. 2013;216(2):175–183.

